# Trends in Breast Pump Prescription Claims: A Nationwide Population-Based Study of Outpatient Statutory Health Insurance Billing Data in Germany, 2011–2024

**DOI:** 10.64898/2026.02.13.26345532

**Authors:** Laura Fischer, Andrea Espinosa Daudí, Zelalem T. Haile, Melissa A. Theurich

**Affiliations:** Chair of Public Health and Health Services Research, Ludwig-Maximilians-Universität München (LMU Munich), Munich, Germany; Pettenkofer School of Public Health, Munich, Germany; German Institute for Drug Use Evaluation (DAPI), Berlin, Germany; Ohio University Heritage College of Osteopathic Medicine, Department of Social Medicine, Dublin, Ohio, USA; Neonatologie der Kinderklinik, Dr. v. Haunerschen Kinderspital und Perinatalzentrum, Ludwig-Maximilians-Universität Klinikum, München, Germany

**Keywords:** breast pumps, medical aids, breastfeeding, lactation, maternal, child health

## Abstract

**Objective:** The objective of this analysis was to explore temporal and regional trends in breast pump prescription claims in outpatient settings in Germany, and to characterize the types of pumps covered.

**Study design:** We conducted a nationwide secondary analysis of outpatient statutory health insurance billing data for breast pump prescriptions from 2011 to 2024, covering nearly 90% of the German population. Billing data from community pharmacies were scaled to full national coverage using regional extrapolation factors and subsequently linked with national and state-level live birth statistics to adjust for birth rates and population size across federal states. A list of breast pumps covered by German national statutory health insurance funds was queried for information on their characteristics.

**Results:** Prescription of electric pumps dominate outpatient statutory health insurance breast pump claims in Germany, with national statutory health insurance funds covering €15.3 million for pump rentals. Manual pumps dispensed through community pharmacies accounted for €27 thousand in 2024. Between 2011 and 2024, electric pump claims increased by a factor of 2.57, rising from 235.4 to 605.2 claims per 1000 infants newly enrolled in statutory health insurance (average annual growth rate 8.24%). Claims varied substantially across federal states but increased overall.

**Conclusions:** This is the first epidemiological analysis of statutory health insurance prescription claims for breast pumps in Germany. We found that electric breast pumps are important medical devices supporting outpatient human milk expression in Germany. Prescription claims appear to be very common and have shown an increase over the past 13 years.

## Introduction

Human milk is widely recognized as the optimal nutrition for infants, supporting healthy development of the infant’s gut microbiome and immune system, while lowering the risk and severity of infectious diseases and offering lifelong health benefits for both mother and child (1, 2). A dose-response relationship has been observed, with longer and more frequent human milk feeding linked to greater protection (3). Human milk is especially critical for premature and low-birthweight infants, where it has been shown to reduce the risk of mortality, necrotizing enterocolitis, late-onset sepsis, retinopathy of prematurity, feeding intolerance, and delayed enteral feeding (4–6). However, mothers of premature and very low birth weight (VLBW) infants are particularly vulnerable to premature breastfeeding cessation (7).

Breast pumps are essential medical aids when direct breastfeeding is not feasible, such as during hospitalization or in cases of clinical breastfeeding difficulties (8). When medically indicated, access to a breast pump can support human milk feeding and improve breastfeeding outcomes. In a large study in the United States, breast pump utilization demonstrated positive effects on breastfeeding duration, reducing the hazard of breastfeeding cessation by 37% and extending breastfeeding by an average of 21 weeks (9). In premature infants, exclusive human milk feeding (whether at breast or expressed) at discharge from the Neonatal Intensive Care Unit (NICU) was associated with higher breastfeeding rates at six months (10). For VLBW infants, early initiation of human milk feeding, often enabled by breast pumps, was the strongest predictor of later volumes of human milk and breastfeeding duration (11).

In Germany, breast pumps are reimbursed by statutory health insurance when they are prescribed by a physician for medical indications affecting the mother or infant. Examples of indications for breast pump use in infants include challenges in coordination of sucking, swallowing, or breathing, prematurity, congenital malformation, muscle weakness, or neurological conditions. Lactating women may require breast pumps for both hypo- or hyperlactation (12) or when they have injuries to the nipple-areola complex. Prescriptions can be issued regardless of infant age and by any treating physician (e.g., general practitioners, gynecologists, pediatricians) (13). With medical indications, statutory health insurances cover outpatient rental or purchase of manual or electric breast pumps. At present, prescriptions for mothers in Germany without a medical indication who pump for work or personal reasons are not covered by statutory health insurances and must pay out-of-pocket. Prescriptions for rental of electric pumps typically last 28 days and up to six months. Community pharmacies usually provide instructions on pump use (14).

In Germany, epidemiological data on breast pump use are lacking and the overall prevalence of milk expression amongst lactating women is unknown. The objective of this study was to provide the first description of temporal and regional trends in outpatient statutory health insurance claims for breast pumps in Germany, and to characterize the types of pumps covered in outpatient settings.

## Methods

In this nationwide population-based study, we conducted a secondary analysis of German statutory health insurance billing data. We explored trends in claims for breast pumps that were prescribed in outpatient settings and billed to German statutory health insurance funds through community pharmacies between 2011 and 2024.

### Data sources

The database of the *German Institute for Drug Use Evaluation (DAPI)* was used to obtain data on statutory health insurance claimed breast pumps. About 89% of the German population (i.e., approximately 75 million people) were covered by statutory health insurance in 2024 (15). The database contains fully anonymized dispensing data from a representative sample of more than 80% (2011 to June 2019) and more than 95% (July 2019 to 2024) of statutory health insurance prescriptions dispensed from community pharmacies in all 16 German federal states. We queried the DAPI database for the following medical aid item numbers of the German *National Association of Statutory Health Insurance Funds*’ medical aids list (*GKV-Hilfsmittelverzeichnis)*:

- *01*.*35*.*01*.*0*.*XXX: Breast Pumps for Manual Use*
- *01*.*35*.*01*.*1*.*XXX: Breast Pumps for Electric Use*

There were no changes in item definition during the analysis period. Birth statistics were obtained from the German *Federal Ministry of Health* (i.e., the nationwide number of infants newly enrolled in German statutory health insurance between 2011-2024) (15). We obtained regional birth statistics from each federal state for each year between 2011-2024 from the German *Federal Statistical Office* (16). The medical aids list of the German *National Association of Statutory Health Insurance Funds* was queried for information on the characteristics of breast pumps that are covered by national statutory health insurance funds (14).

The primary outcome was the annual number of outpatient breast pump prescription claims per 1000 infants. Additionally, types of pumps covered in outpatient settings were categorized in terms of their characteristics.

### Statistical Analysis

Using regional extrapolation factors, billing data was scaled up to represent 100% of community pharmacies dispenses (expenses to the statutory health insurances). Up to June 2019, these factors were calculated by dividing the total number of community pharmacies in a region by the number of pharmacies covered by the DAPI database in the respective region. The year was used as the time unit of data aggregation. We linked the billing data with national and state-level birth statistics over time to adjust for variations in birth rates and differences in population size between the states. For national estimates, we used live birth statistics from the statutory health-insured population to calculate “claims per 1000 infants newly enrolled in German statutory health insurance”. These administrative data were only available at the national level. For state-level estimates, we calculated “claims per 1000 live births” using general population birth statistics instead. Because the state-level denominator includes both statutory and privately insured births, these rates can be interpreted as conservative estimates of the statutory health insurance-based rates and may vary by region depending on private health insurance coverage.

Data analysis was exploratory thus no hypotheses were prespecified. We descriptively analyzed the number of breast pump claims in the analysis period, 2011-2024. We further conducted a trend analysis using the Mann-Kendall trend test to statistically assess the direction of the trend of claims over time. Due to the low number of prescriptions, regional analyses were not conducted for manual pumps. We applied a local regression (“loess”) to fit smooth curves to the empirical data (17). All analyses and data visualizations were performed using R Software 4.1.0 (18).

In order to describe the characteristics of breast pumps available with coverage from the German statutory health insurances, descriptive data was extracted and categorized from the medical aids list of the German *National Association of Statutory Health Insurance Funds* (14).

### Study Protocol and Reporting

We publicly registered our study protocol on the Open Science Framework (19). We originally planned to report on both breast pumps and pumping accessories, such as flanges. However, data on pumping accessories could not be differentiated from other suction equipment (unrelated to breast pumps) and were thus excluded from this analysis. We report no further deviations from the study protocol. The data and R code can be made available by the authors upon reasonable request. We report our findings according to the REporting of studies Conducted using Observational Routinely collected health Data (RECORD) statement (20).

## Results

In 2024, German national statutory health insurance funds covered costs of €15.3 million for electric pumps and €27 thousand for manual pumps that were dispensed through community pharmacies. Electric pump claims increased in absolute numbers by a factor of 2.52 between 2011-2024, rising from n=134,769 in 2011 to n=339,951 in 2024, with a peak observed in 2020 at n=405,612 claims (see **Figure 1A**). Adjusting for annual variations in birth rates, electric pump claims per 1000 infants newly enrolled in statutory health insurance increased by a factor of 2.57, from 235.4 in 2011 to 605.2 in 2024 (see **Figure 1B**). This corresponds to an average annual growth rate of 8.24%.

**Figure 1:**
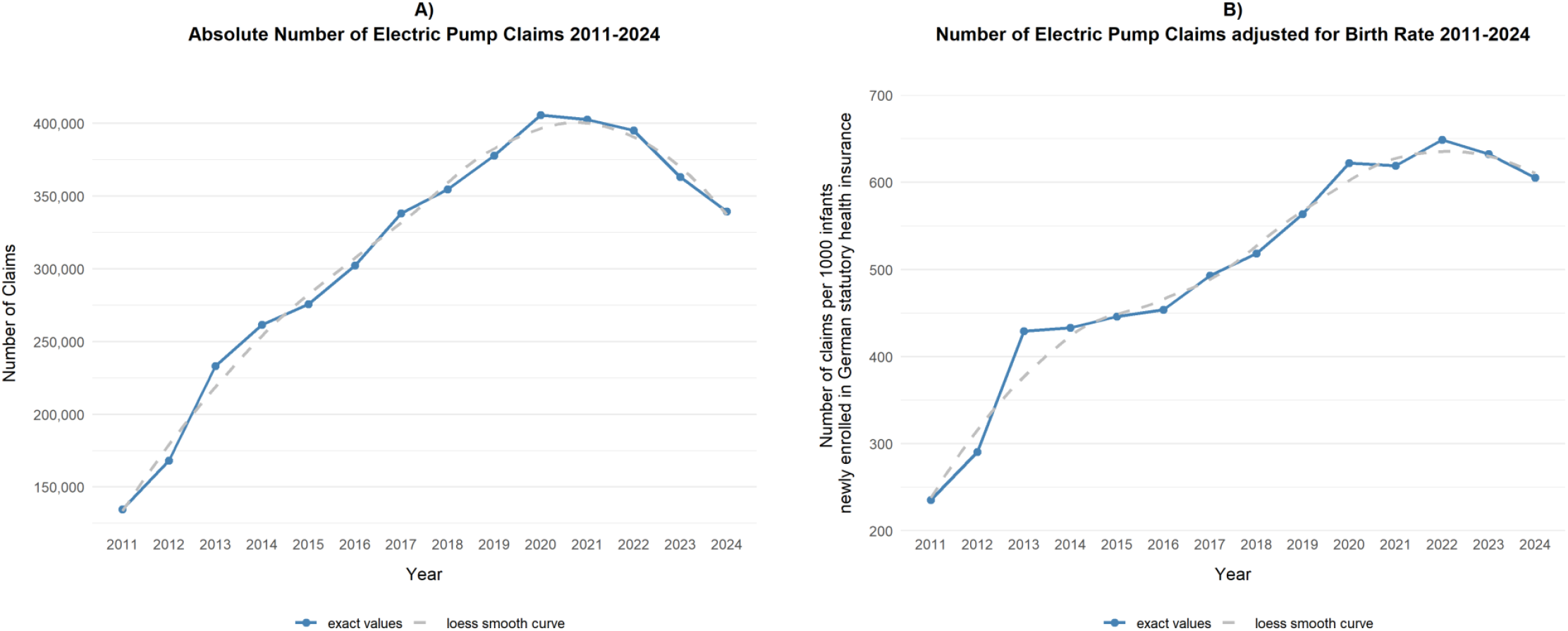
Trend in electric pump health insurance claims 2011-2024.

Manual pump claims decreased by 80.9% from 5.18 to 0.99 claims per 1000 infants, with an average annual decrease of 6.90%. The absolute number of manual pump claims decreased by a factor of 5.34 from n=2966 in 2011 to n=555 claims in 2024, see **Figure 2**.

**Figure 2:**
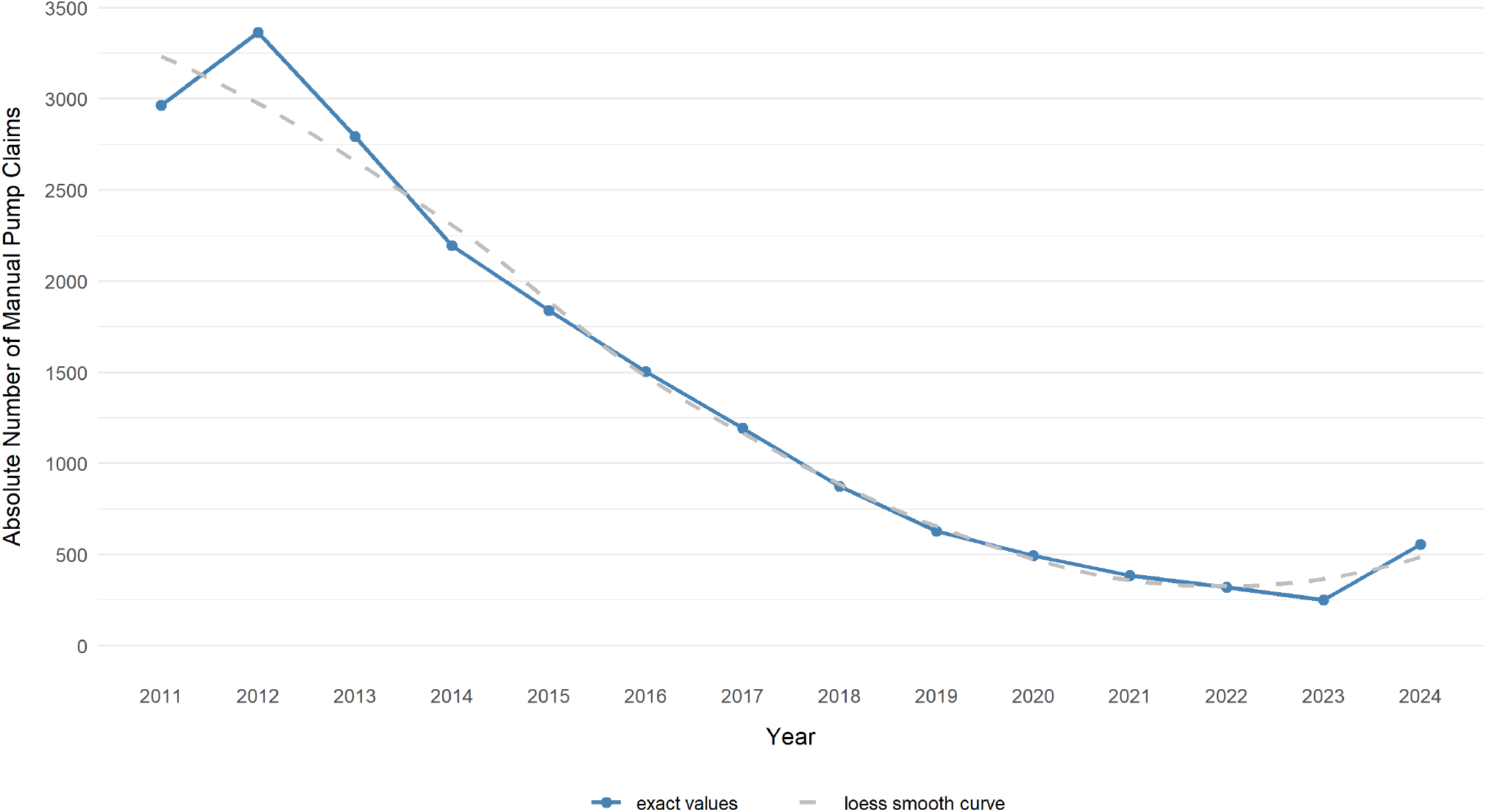
Trend in manual pump health insurance claims 2011-2024.

The Mann-Kendall trend test revealed a statistically significant upward trend in electric pump claims per 1000 infants from 2011 to 2024 (τ = 0.87, *p* < 0.001). This suggests a consistent increase in electric pump claims over the analysis period. Manual pump prescriptions showed a statistically significant downward trend over the same time period (τ = −0.87, *p* < 0.001). There is a strong inverse correlation between the two variables (r = −0.93). However, as these are aggregate time-series trends, this correlation should be interpreted cautiously and not as evidence of a direct substitution effect at the individual prescriber or patient level.

### Regional differences in electric pump health insurance claims

There were regional differences in the number of electric pump claims across German federal states. Variations across German states over time are visualized in **Figure 3**, showing that the number of claims increased in each federal state between 2011 and 2024. There are strong variations in annual growth rates with some defined jumps at different years across the states. Saxony had the highest number of claims per 1000 live births consistently.

**Figure 3.**
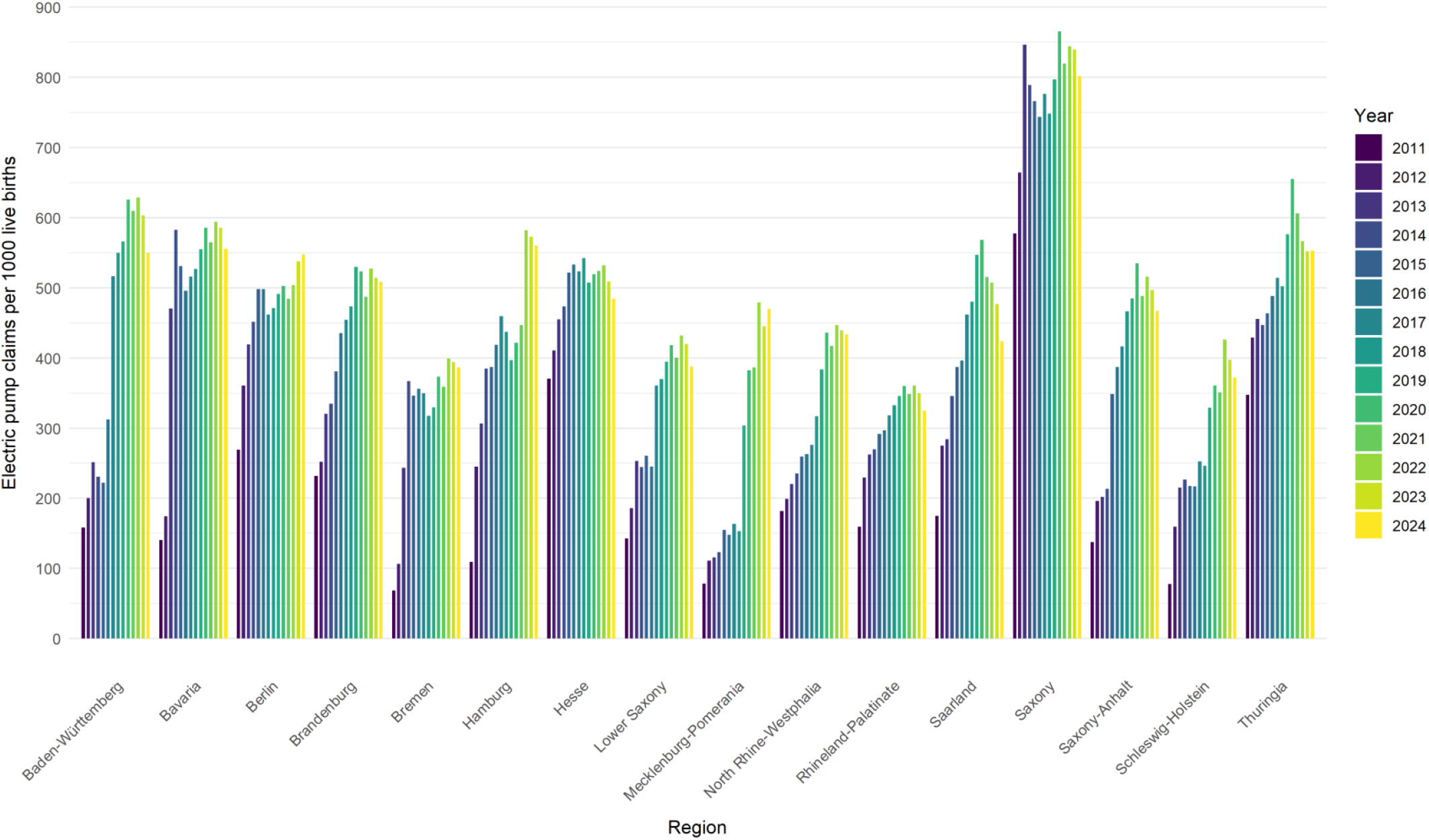
Regional trends of electric pump claims at the expense of statutory health insurances across German states per 1000 live births over time, 2011-2024.

Regional variations of electric pump claims at the expense of statutory health insurances are further described cartographically in **Figure 4:** In 2024, Saxony recorded the highest number of claims per 1000 live births (801.7), while Rhineland-Palatinate had the lowest (325.2), see **Figure 4A**. The total change per 1000 live births between 2011 and 2024 was largest in Mecklenburg-Western Pomerania (+ 500.2%) and Bremen (+ 464.1%), where there were around five times more claims in 2024 than in 2011, and smallest in Hesse (+ 30.7%), Saxony (+ 38.9 %), and Thuringia (+ 59.1 %), see **Figure 4B**. The states with the smallest growth were the states with the highest number of claims per 1000 live births already in 2011. All exact values, including the regional Mann-Kendall trend statistics, are provided in **Table 1**.

**Table 1.**
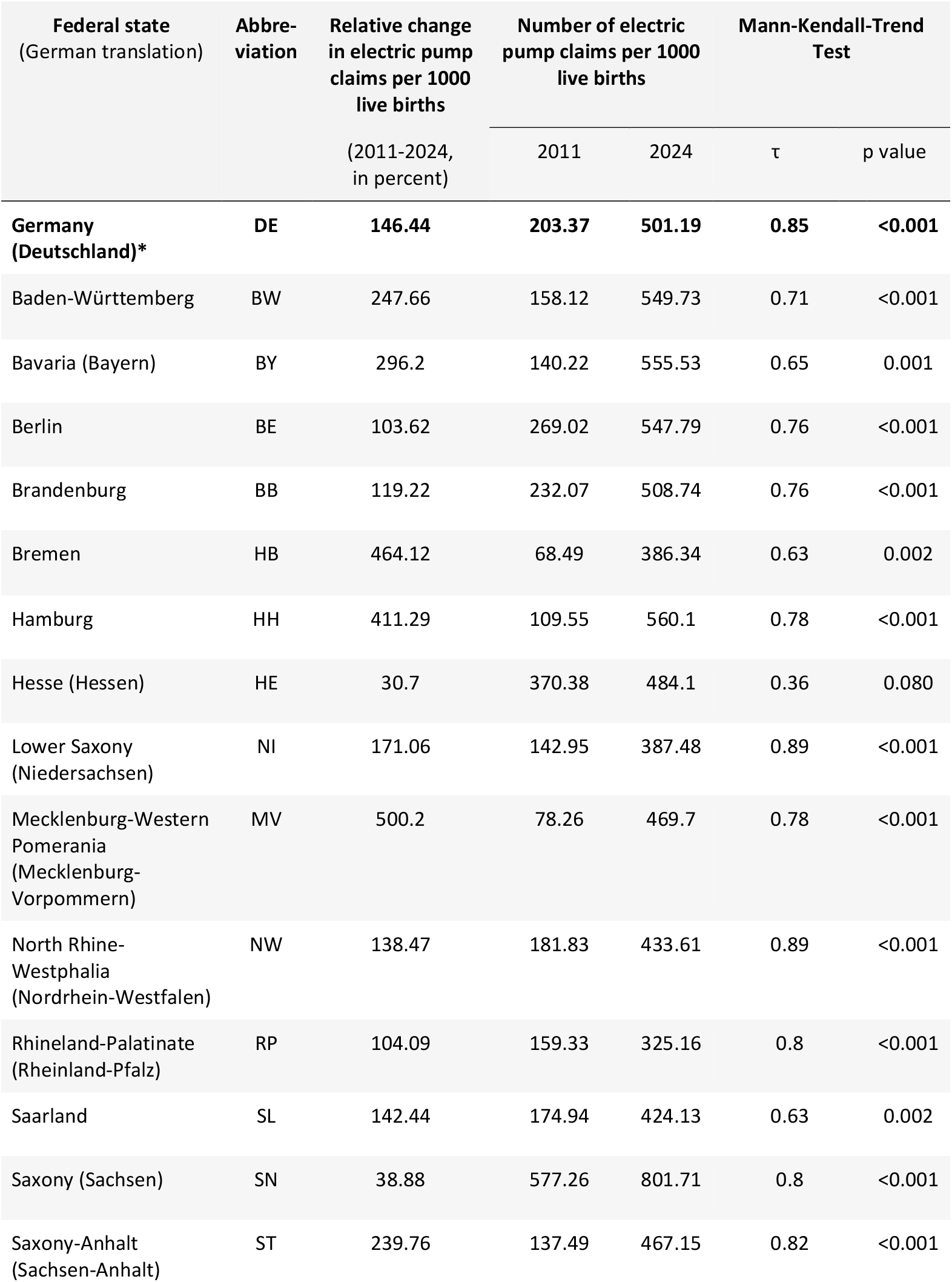
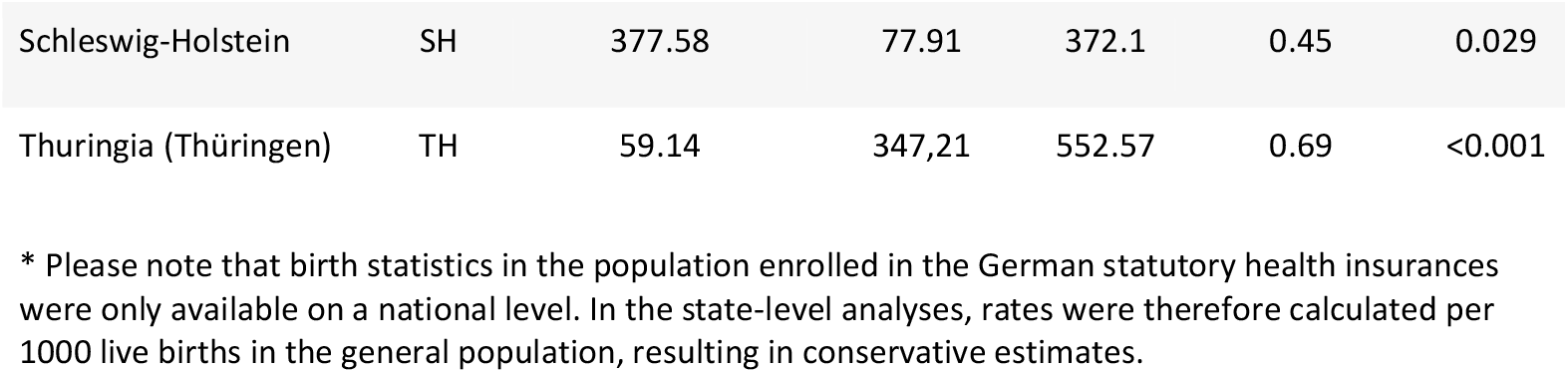
Trends in electric breast pumps claimed to the German statutory health insurance between 2011 and 2024 in the German federal states.

**Figure 4.**
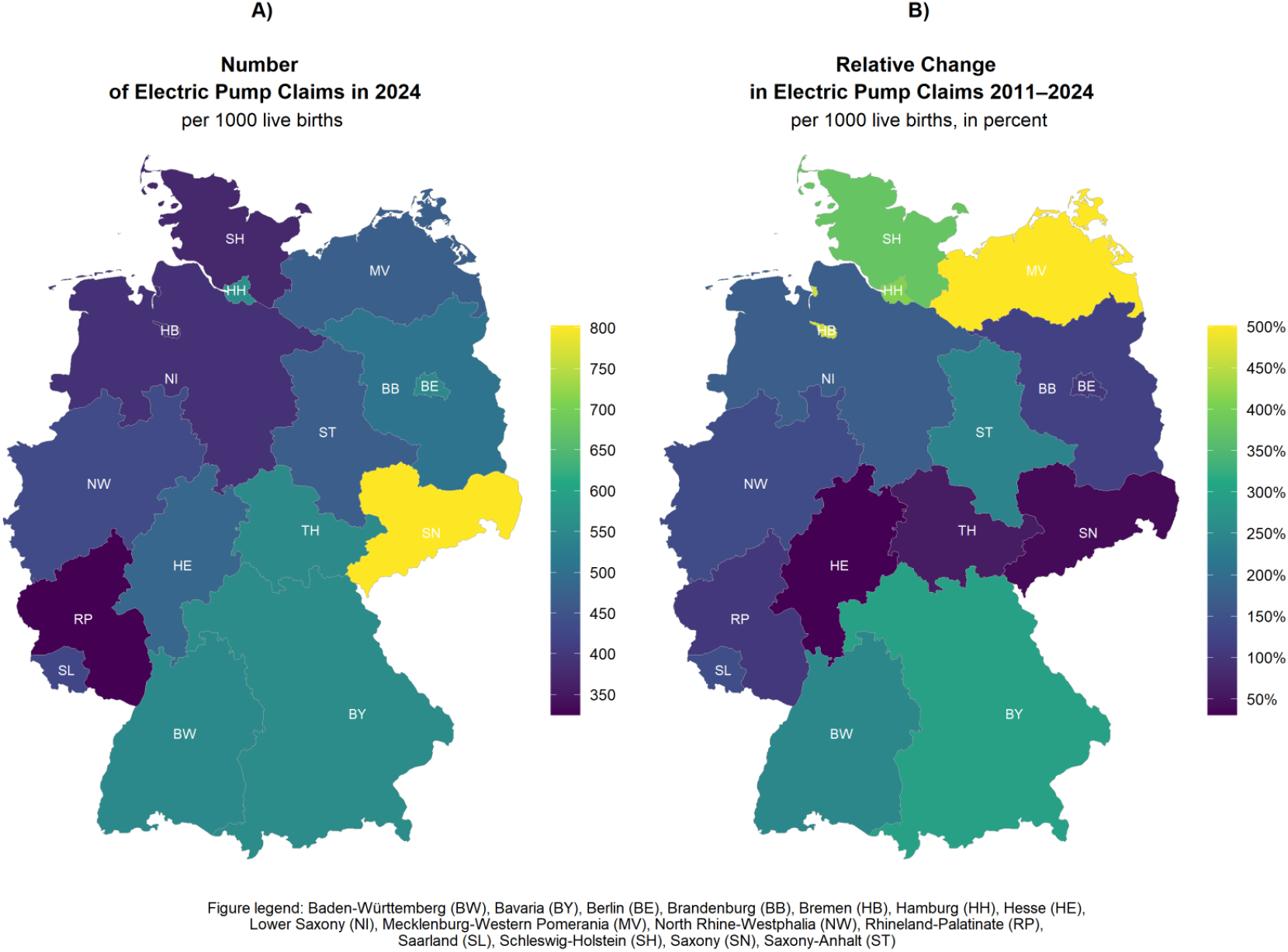
Regional differences in electric pump claims at the expense of statutory health insurances across Germany.

### Characteristics of breast pumps covered by German statutory health insurance

As of January 2025, there were 20 breast pumps covered by German national statutory health insurance funds (14). Most pumps were electric (90%, n=18); only two pumps were manual. The accessories for two of the electric pumps could also be used manually. Most electric pumps (83%, n=15) were double-sided and 39% (n=7) had an integrated battery. The majority (72%, n=13) reported having an adjustable interval cycle for suction with an average range of 34 to 70 cycles per minute (min: 20, max: 120). The maximum suction strength was on average 250 mmHg (min: 240 mmHg, max: 300 mmHg). Most pump manufacturers covered by German national statutory health insurance funds were headquartered in Germany (6 manufacturers; 15 pumps), followed by the US (1 manufacturer; 3 pumps), Turkey and France (1 manufacturer and 1 pump each).

## Discussion

This nationwide analysis of German statutory health insurance billing data provides the first insights into the trends of outpatient utilization of breast pumps in Germany, reflecting use of breast pump among mothers who obtained a pump due to a medical indication in themselves or their infants. Between 2011 and 2024, claims for electric pumps grew by a factor of 2.57, corresponding to an annual average growth rate of 8.24%. Our results show a clear preference for electric pumps over manual pumps, while prescriptions for manual pumps were minimal at the national level. This is also reflected in the list of medical aids that are covered by German national statutory health insurance funds, in which electric pumps constitute the vast majority of devices. The number of electric pump claims per 1000 live births varied substantially across German federal states but showed an overall increasing trend. No consistent east-west or north-south difference was observed.

To our knowledge, there were no changes in health care policies or coverage that would explain the overall growth or the regional differences in breast pump prescription claims at the expense of statutory health insurances in Germany. However, our results are in line with reports from other countries demonstrating increasing rates of human milk expression. For example, a study from Australia showed that the proportion of mothers who had expressed human milk within the first six weeks after birth had almost doubled (from 38% to 69%) between 1993 and 2003 (21). A cohort study from Singapore showed an increase in expressed milk feeding in more recent birth years (2000-2001: 8.6% vs. 2006-2008: 18.0%) (22). In a cross-sectional study including 19,780 women in the United States, most breastfeeding women (90.6%) reported having used a pump (9).

In our analysis, we observed 605.2 electric pump claims at the expense of German statutory health insurances per 1000 infants newly enrolled in German statutory health insurance in 2024. Since the aggregated data do not indicate whether individual mother-infant dyads received follow-up prescriptions, which we believe to be common in clinical practice, we considered two scenarios to estimate prevalence. Under the assumption that no mother-infant dyad used a pump for longer than 28 days, 605.2 pump claims per 1000 infants would imply that electric pumps were used to provide human milk to roughly 6 in 10 infants (60% of infants; upper-bound estimate). In a more realistic scenario, in which each mother-infant dyad rented an electric pump for three months (requiring three 28-day prescriptions), prescribed electric pumps would have been used to support human milk provision for about 2 in 10 infants (20% of infants; lower-bound estimate).

The increased use of breast pumps internationally coincides with market research that shows a growing breast pump market (23). Empirical literature suggests an overall increase in innovation around breast pump technology since 2010 due to improved insurance coverage in the United States (24) and that breast pumps are increasingly efficient, safe, comfortable and convenient to use (25, 26).

The observed growth in breast pump prescription claims at the expense of German statutory health insurances may reflect increasing use among lactating women seeking support for clinical breastfeeding difficulties, as well as greater awareness among physicians regarding pump prescription. Literature further suggests that the observed increase in breast pump prescriptions may partly reflect marketing practices that portray pumping as necessary or natural (27, 28). Such messaging may also influence healthcare providers’ prescribing practices and contribute to the rise in prescription claims. Although breast pumps can support human milk feeding, they are not indicated or required for all breastfeeding dyads (8). This underscores the importance of healthcare professionals and public health stakeholders providing reliable information on both the health benefits and risks of breast pump use to lactating women (8).

Despite public health recommendations (29, 30), breastfeeding rates in Germany remain low with only 19% of infants exclusively breastfeeding at six months, and only 57% being breastfed at all (31). Breast pumps are a key technology for managing breastfeeding difficulties and supporting families in achieving human milk feeding goals when direct breastfeeding is not, or only partially, possible in both healthy and vulnerable infants (9–11). This work may be informative to future research and health policy in Germany and Europe. Future epidemiological studies monitoring breastfeeding rates should consider the potential impact of increasing breast pump use when interpreting their results. The regional differences in breast pump prescription claims at the expense of German statutory health insurances observed in our analysis may reflect variations in prescribing practices, clinical breastfeeding difficulties or attitudes toward pumping. This calls for future in-depth research to further better understand regional determinants of infant feeding choices, human milk expression and breastfeeding practices in Germany.

### Strengths

In Germany, nearly 90 % of the German population (which is approximately 75 million people) was covered by statutory health insurance in 2024. Therefore, the major strength of this study is its comprehensive and valid statistical basis, consisting of a large population-based representative sample of more than 80% (2011 to June 2019) and more than 95% (from July 2019) of the claims to the German statutory health insurance funds from community pharmacies in all 16 German federal states. We were able to describe health insurance prescription claims for breast pumps over a time period of 13 years, as well as regional differences at the state-level.

## Data Availability

All data produced in the present study are available upon reasonable request to the authors.

## Limitations

This was a secondary analysis of routine health insurance data and therefore has some limitations. Our data are limited to outpatient billings from community pharmacies to the German national statutory health insurance funds. They therefore do not include pumps covered by private health insurances (which enroll approximately 10% of the population), pumps paid out-of-pocket, or pumps distributed in in-patient settings, by other outpatient vendors such as medical supply stores, or other private health providers like midwives or lactation consultants. One claim corresponds to a single prescription for a duration of up to 28 days. Based on the available aggregated data, we cannot know if single mother-infant dyads had multiple follow-up prescriptions, which we believe to be common in clinical practice, and how many dyads were ultimately supported. Furthermore, the database includes all claims to the German statutory health insurances and not only reimbursed claims.

Birth statistics in the population enrolled in the German statutory health insurances were only available on a national level (15). In the state-level analyses, rates were therefore calculated per 1000 live births in the general population. This results in conservative estimates as only 89% of Germans are enrolled in statutory health insurance (15). Since private insurance coverage varies across regions, underestimation is likely non-uniform, limiting the direct quantitative comparisons of regional rates. We could not obtain any information on the regional differences in coverage of newborns by German statutory health insurances. Multiple births are unlikely to meaningfully affect our assumption of one breast pump per child, as twins accounted for only 1.84% of newborns in 2019 and their rates do not substantially vary by region (32). We therefore did not control twin births in our analysis. Finally, because the Mann-Kendall test does not account for autocorrelation in annual time-series data, our trend p-values should be viewed as descriptive indicators of monotonic change rather than formal inferential tests.

## Ethics statement

This study used anonymized billing data. No individual patient-level information was accessed, and ethical approval and informed consent were not required. Clearance for working with the data was given by the LMU Munich data protection officer.

## Authors Contributions

LF: Conceptualization; Data curation; Methodology; Formal analysis; Visualization; Writing - original draft; and Writing - review & editing

AED: Data curation; Writing - review & editing

ZH: Methodology; Writing - review & editing;

MT: Conceptualization; Methodology; Writing - original draft; and Writing - review & editing; Project administration; Supervision; Funding acquisition

